# Factors Affecting Medication Adherence Among patient with Schizophrenia: A Literature Review

**DOI:** 10.1101/2022.01.12.22269187

**Authors:** Faizatur Rohmi, Moses Glorino Rumambo Pandin

## Abstract

This review aimed to summarize empirical evidence concerning to the factors relating to medication adherence among patients with schizophrenia. A comprehensive search was implemented to recruit articles which met the present eligibility criteria. Twenty articles were included, all of which were quantitative studies. Greater awareness of illness (insight), previous history of medication adherence, positive attitude toward medication, types of atypical antipsychotics, less severe psychotic symptoms, and social support, self-efficacy, general health status, gender men, lower socioeconomic status, living alone, length of illness, drug abuse, education level, severity of illness for example being in the acute phase, participating in mental health services, health facilities, marital status, receiving monotherapy were identified as factors of medication adherence. Implications to clinical practice include providing psychoeducation to patients and family by increasing their knowledge about illness and medication.

## INTRODUCTION

Schizophrenia is a psychotic disorder which consists of a range of features and affects 20 million people worldwide which is characterized by distortions in thinking, perception, emotion, language, and behavior as well as an inability to make interpersonal relationships with other people in time, place and environment (American Psychiatric Association, 2013; Townsend & Morgan, 2018; Videbeck, 2020; WHO, 2019). Based on the results of the 2018 Riskesdas in Indonesia, it was stated that the prevalence of schizophrenia in the 2013-2018 period was 7 per mile (Kemenkes RI, 2018). Based on years of life with disability (YLDS) mental disorders have a contributor of 13.5%, with indicator coverage of people with mental disorders receiving treatment and not being neglected reaching 37.47% in East Java (Kemenkes RI, 2019).

Schizophrenia requires long-term treatment which usually comprises antipsychotics and psychosocial interventions (Townsend, 2011; Videbeck, 2020). Antipsychotics comprise two classes: typical and atypical (Meltzer, 2013). Despite the available treatments, schizophrenia is also marked by frequent relapses and remission (American Psychiatric Association, 2013). The lack of insight about one’s mental condition is common among individuals with schizophrenia and may also impede adherence to medication (American Psychiatric Association, 2013). Adherence is a complex phenomenon and is very difficult to detect when people with psychotic disorders are not undergoing hospitalization (Leijala et al., 2021). Various studies stated that this non-adherencee is one of the reasons a person can relapse, return to the hospital and this will have an impact on a higher treatment costs (Abdisa et al., 2020); (Chai et al., 2021) Various precipitating factors cause non-adherencee to treatment, but the main factor is not yet known with certainty. Based on several research results, 19.1% did not adhere to treatment (Şahin Altun et al., 2021).

The problem of adherence to medication is not a problem unique to schizophrenia. People with conditions such as obstructive pulmonary disease, diabetes, sexually transmitted diseases, cardiovascular disease, chronic pain, and other mental disorders also have problems adhering to medication (Ingersoll & Cohen, 2008). Adherence is defined as “the extent to which a person’s behavior in taking medication, following a diet, and/or making lifestyle changes, is in accordance with agreed recommendations from health care providers” (WHO, 2019). Therefore, medication adherence in this review was determined as a behavior that must be followed by prescription drugs (antipsychotics) according to doctor’s orders.

Adherence can have an immediate impact. Drug nonadherence to psychotic medication can worsen psychiatric symptoms (Perkins, 1999) and social functioning (Perkins, 1999) and can lead to suicide (Perkins, 1999). Return costs for nonadherence to hospital admissions among patients with schizophrenia ranged from US$1392 million to $1826 million in the United States in 2005 (Perkins, 1999) Therefore, the impact not only affects individuals, but also society.

It is important to understand factors associated with medication adherence among people with schizophrenia. As such, this literature review was conducted to summarize empirical evidence concerning factors relating to medication review.

## METHODS

Comprehensive literature search for published studies was first conducted by using the following electronic databases: SCOPUS, SCIENCE DIRECT, SAGE, PsychINFO, and PubMed. Next, reference lists of retrieved articles were also scanned through to retrieve additional articles. The inclusion criteria were studies that (a) were written in English in the last 5 years (2019–2021), (b) utilized quantitative research designs, (c) recruited adult participants with schizophrenia aged between 18 to 65 years, and (d) examined factors relating to medication adherence. The keywords used in the search included “medication”, “adherence”, “non-adherence”, “adherencee”, “non-adherencee”, “antipsychotic”, “neuroleptic”, “schizophrenia”, “psychosis”, “predictor”, “reason”, “factor”, “variable”, “determinant”, “element”, “component”, “aspect”, “belief”, “attitude”, “influence”, and “effect”. This review excluded studies that conducted on participants who had criminal records, as they might not represent the general schizophrenia population. All included studies were also assessed for methodological quality by using the Joanna Briggs Institute (JBI) Critical Appraisal Instrument (JBI, 2014).

Articles obtained in the search process are then selected based on inclusion and exclusion criteria that shown in table 1:

**Tabel 1.** Inclusion and exclusion criteria with PICOS.

| Criteria | Inclusion | Exclusion |
| --- | --- | --- |
| Population | schizophrenia | Not shizophrenia |
| Intervention | There are no intervention | There are no criteria exclusion |
| Comparisons | There are no comparison<br>There are no criteria exclusion | There are no comparison<br>There are no criteria exclusion |
| Outcome | Factors of therapy adherencee in patients with schizophrenia | Not relevant to therapy adherencee factors in people with schizophrenia |
| Study Type | Cross sectional, longitudinal study | mixed method, a quasi experimental design, clinical trial or randomized controlled trial, Systematic or literature reviews, qualitative research |
| Publication Type | Peer reviewed original studies<br>Non peer-reviewed studies | Peer reviewed original studies<br>Non peer-reviewed studies |
| Publication Years | 2019 – 2021 | Pre 2019 |
| Languange | English | Non English Languange |

**Table 2.**
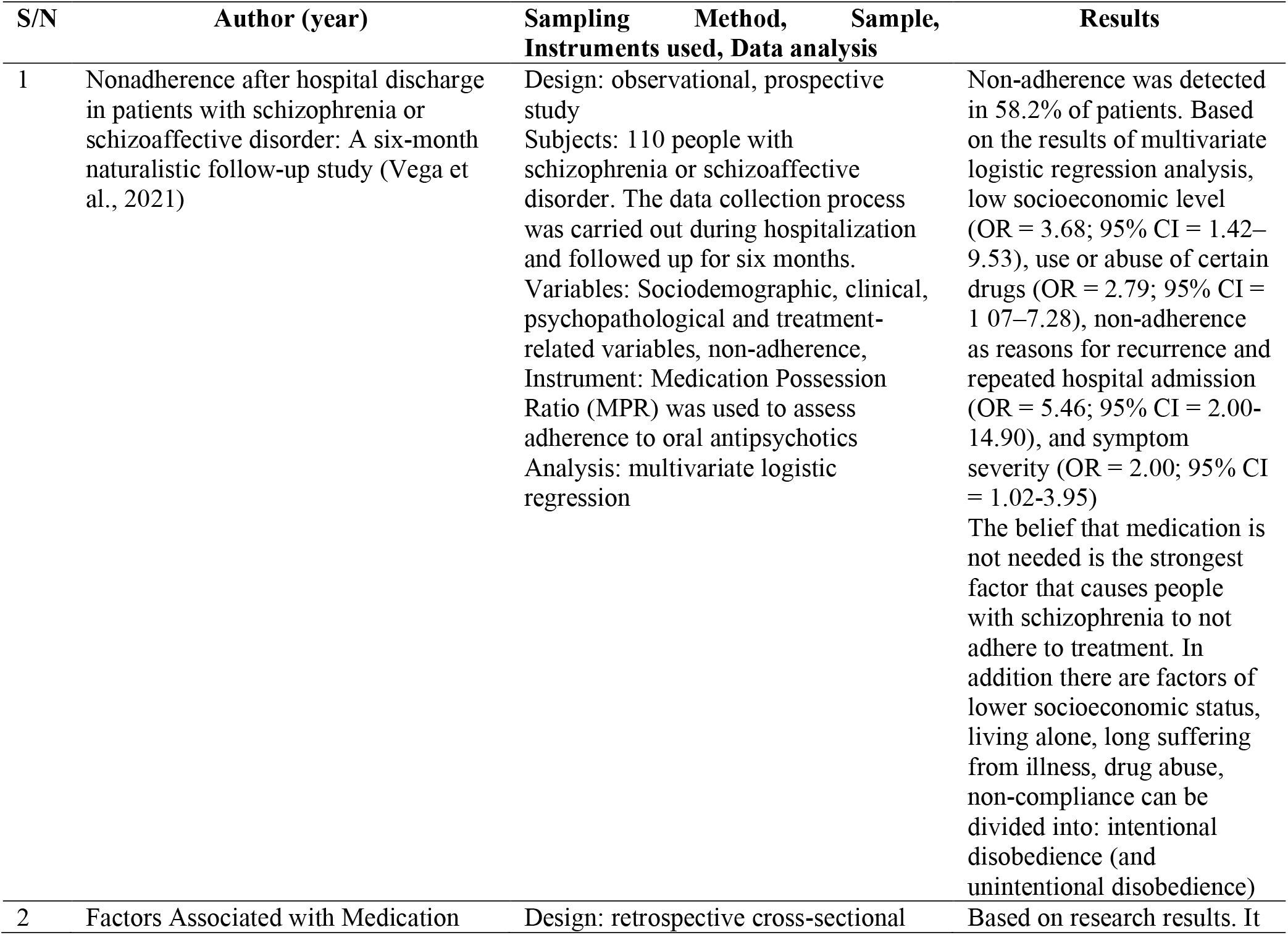
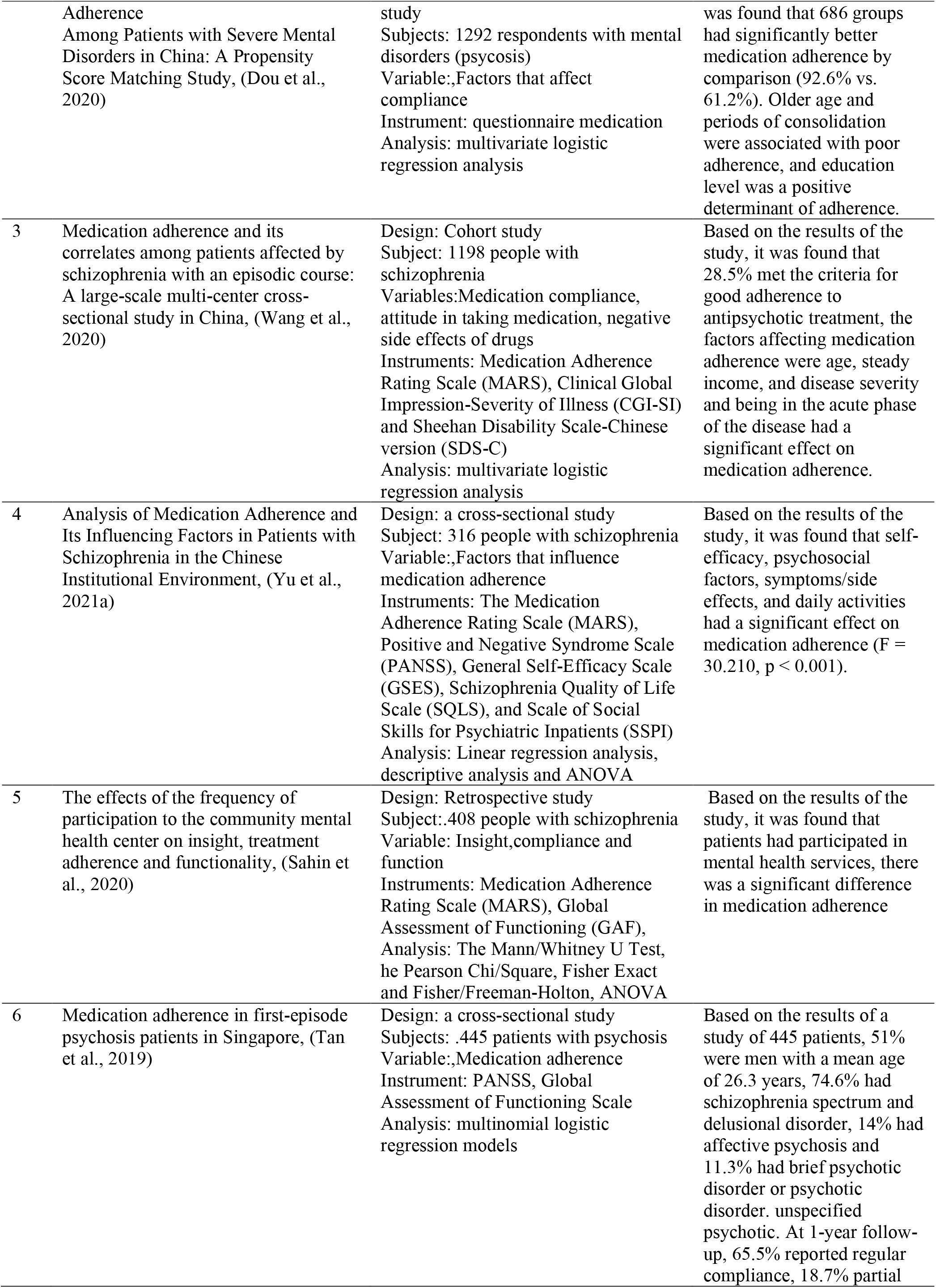
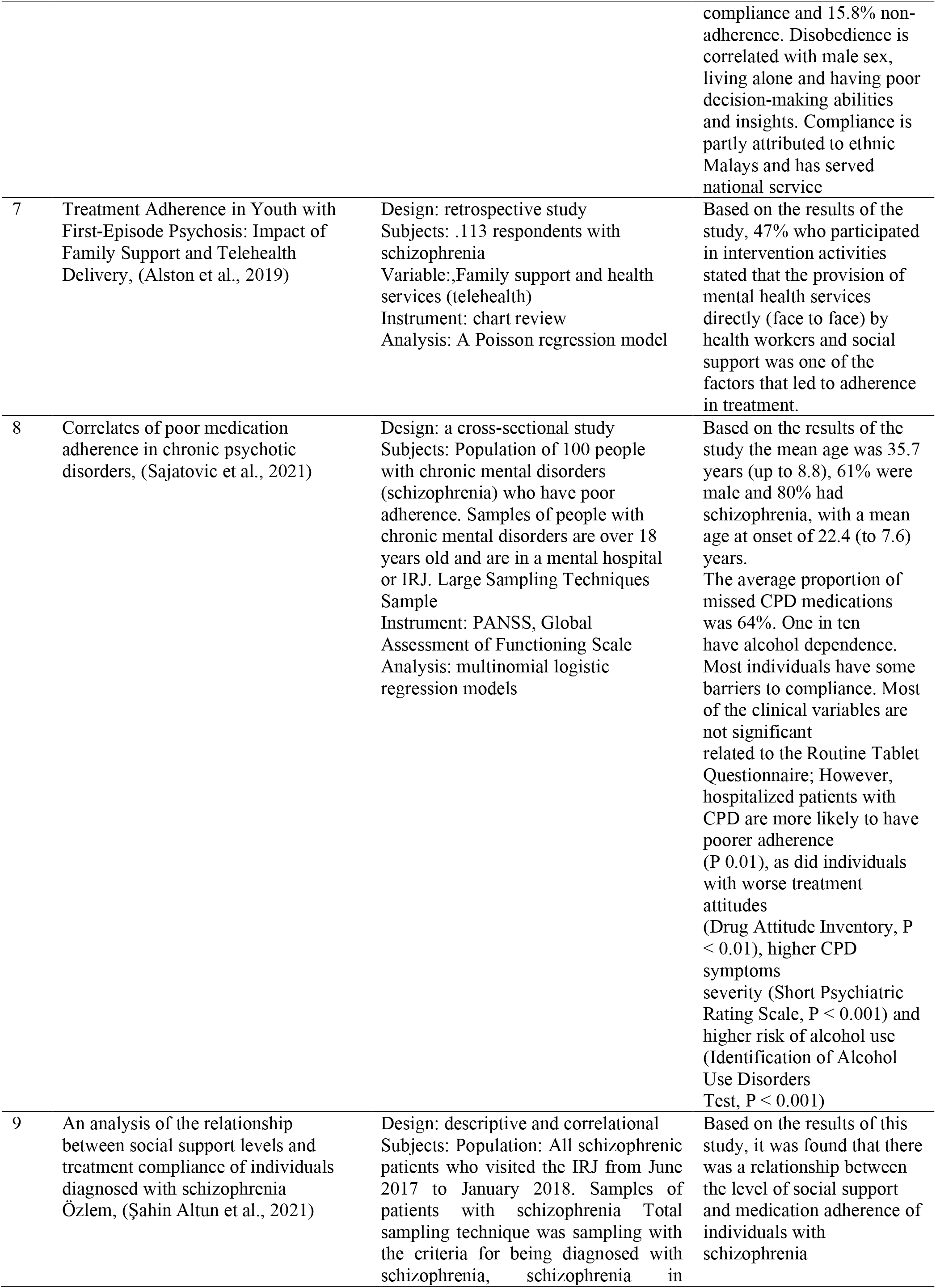
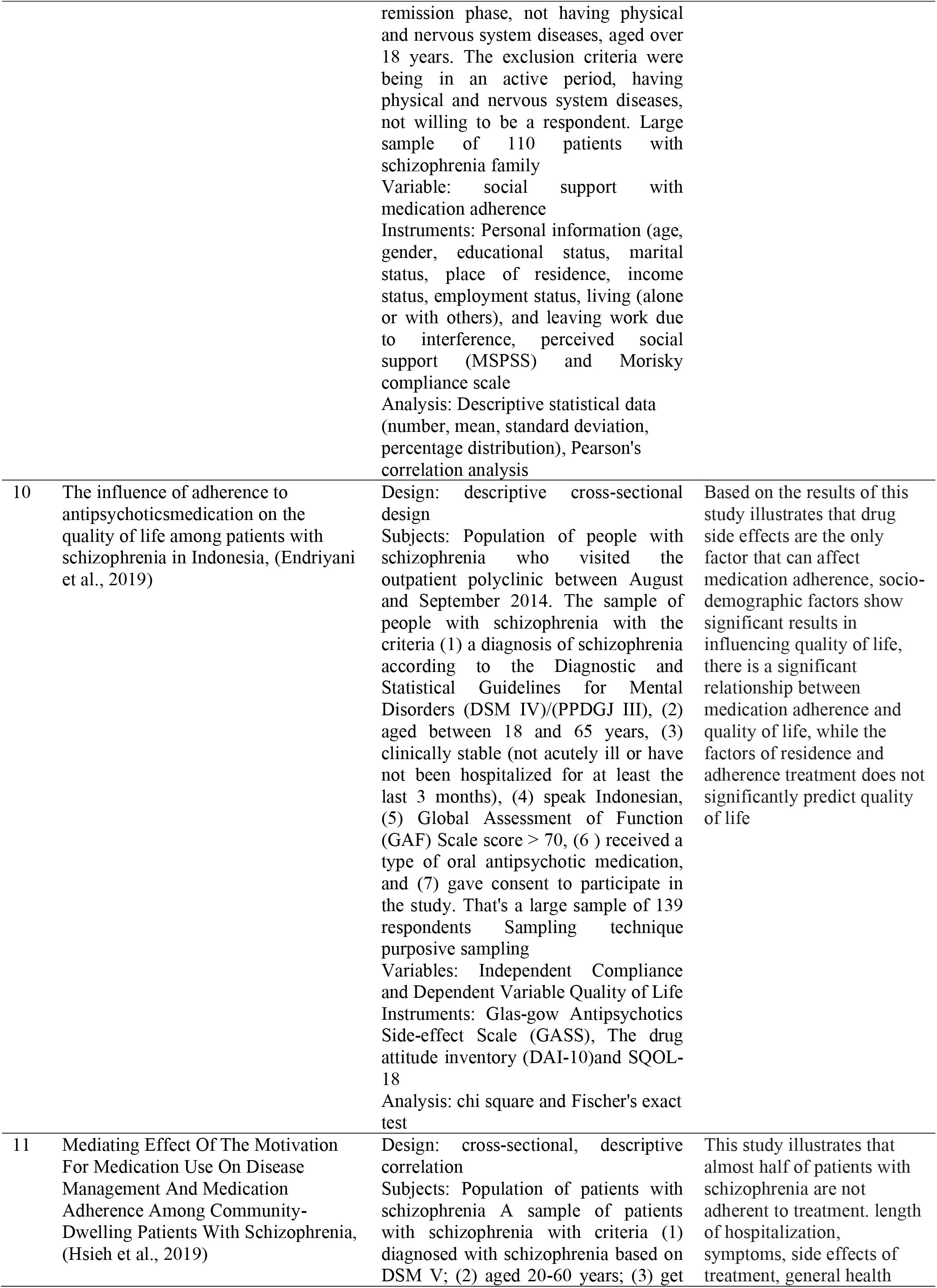
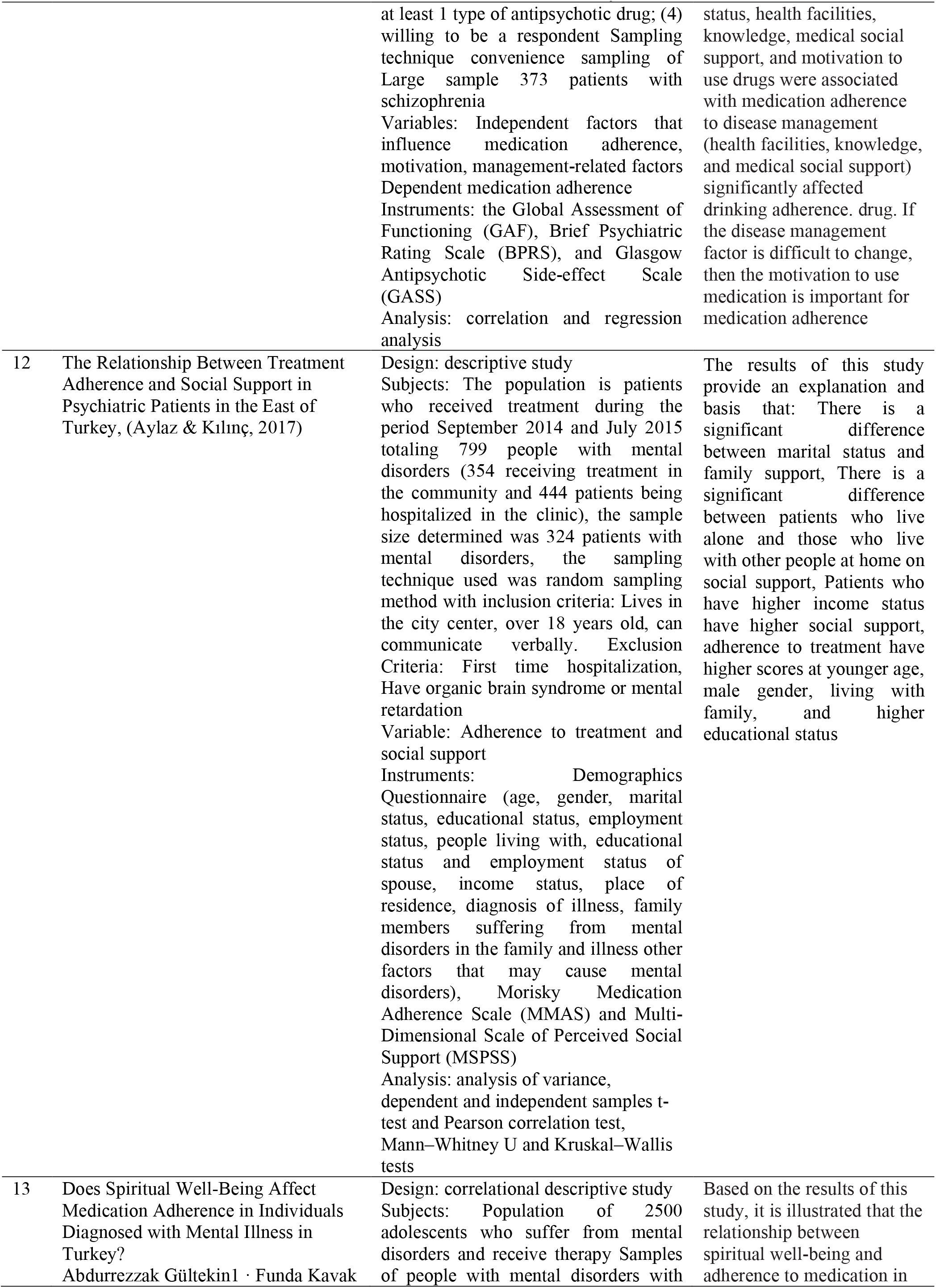
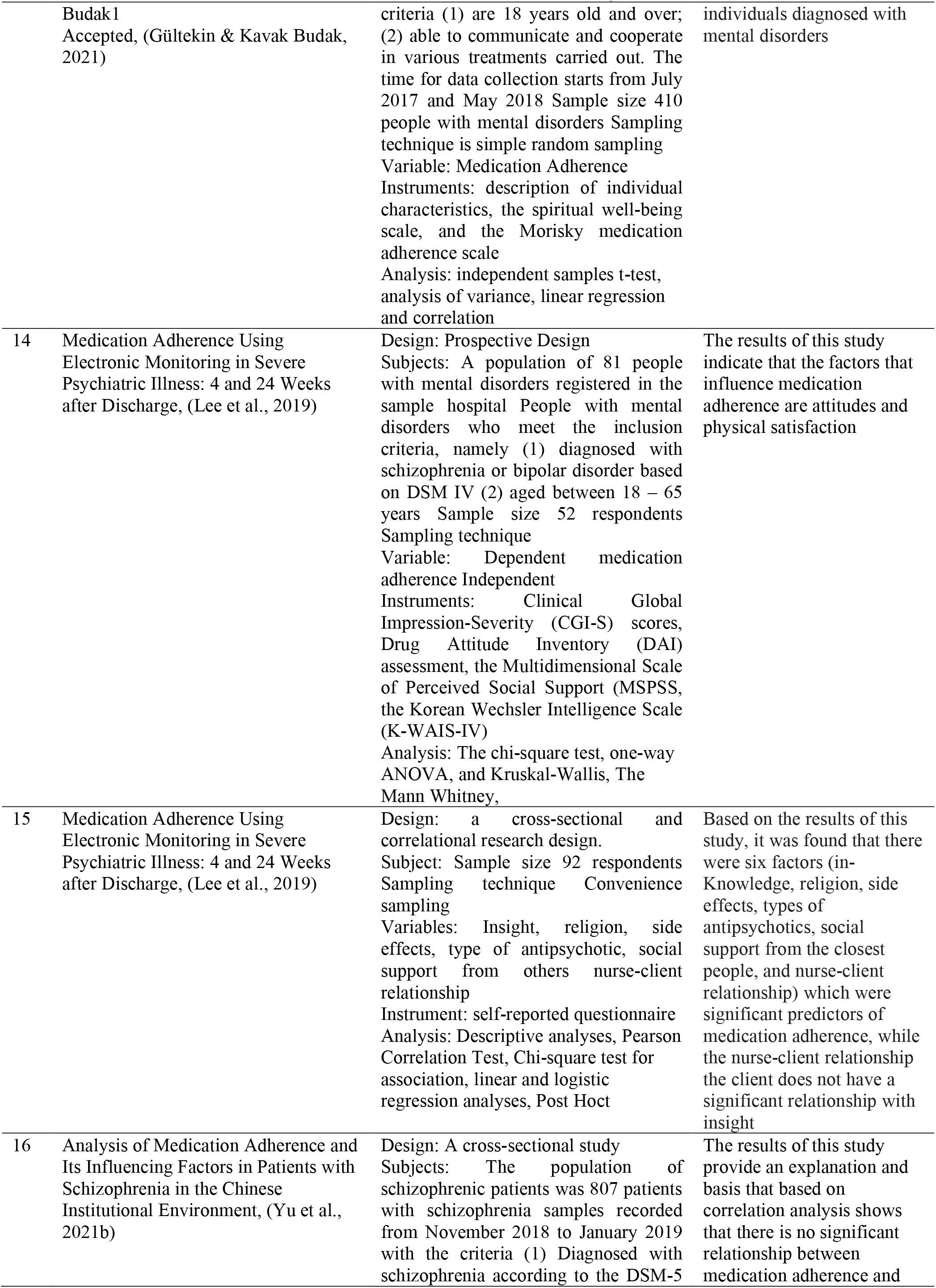
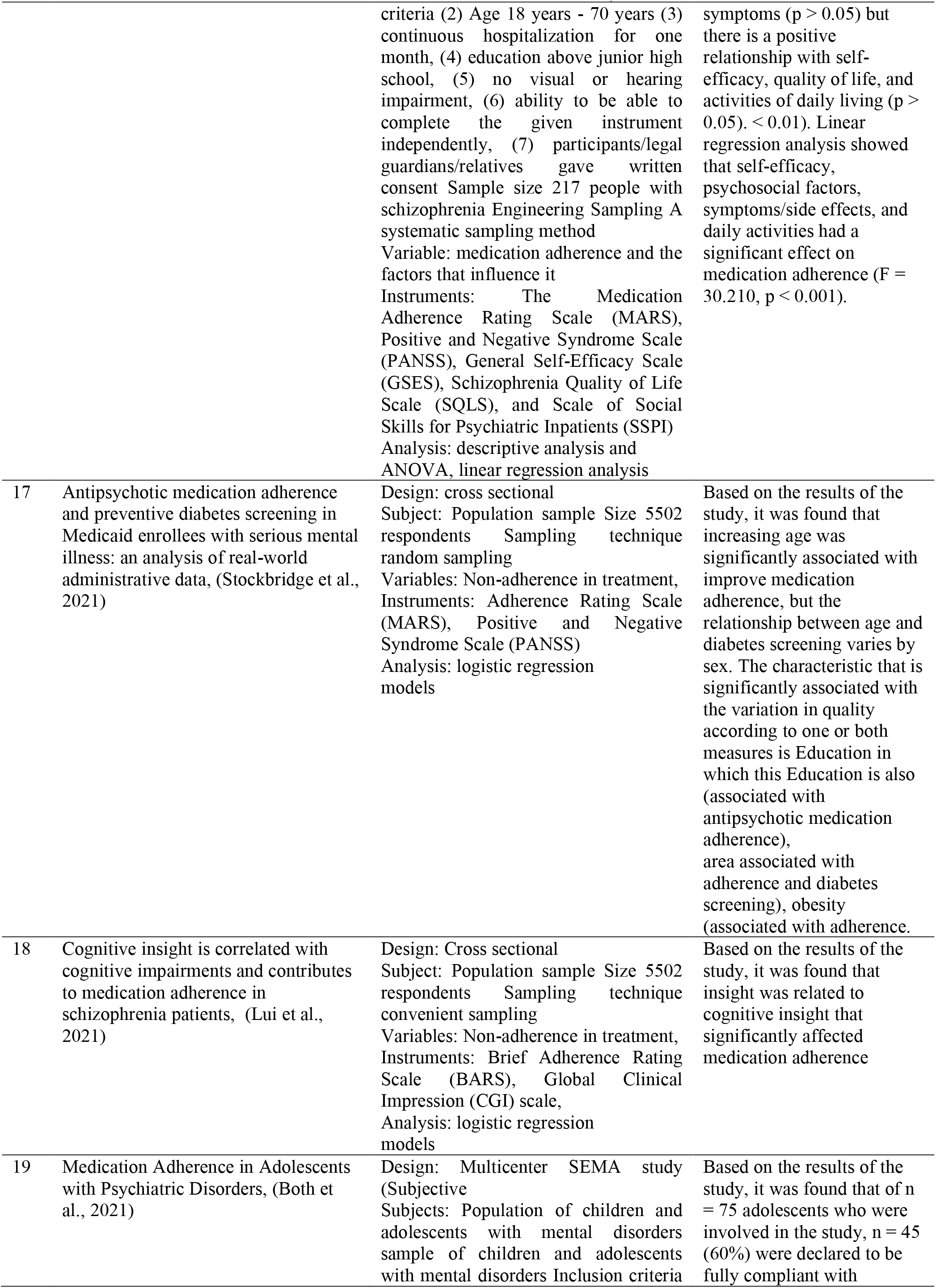
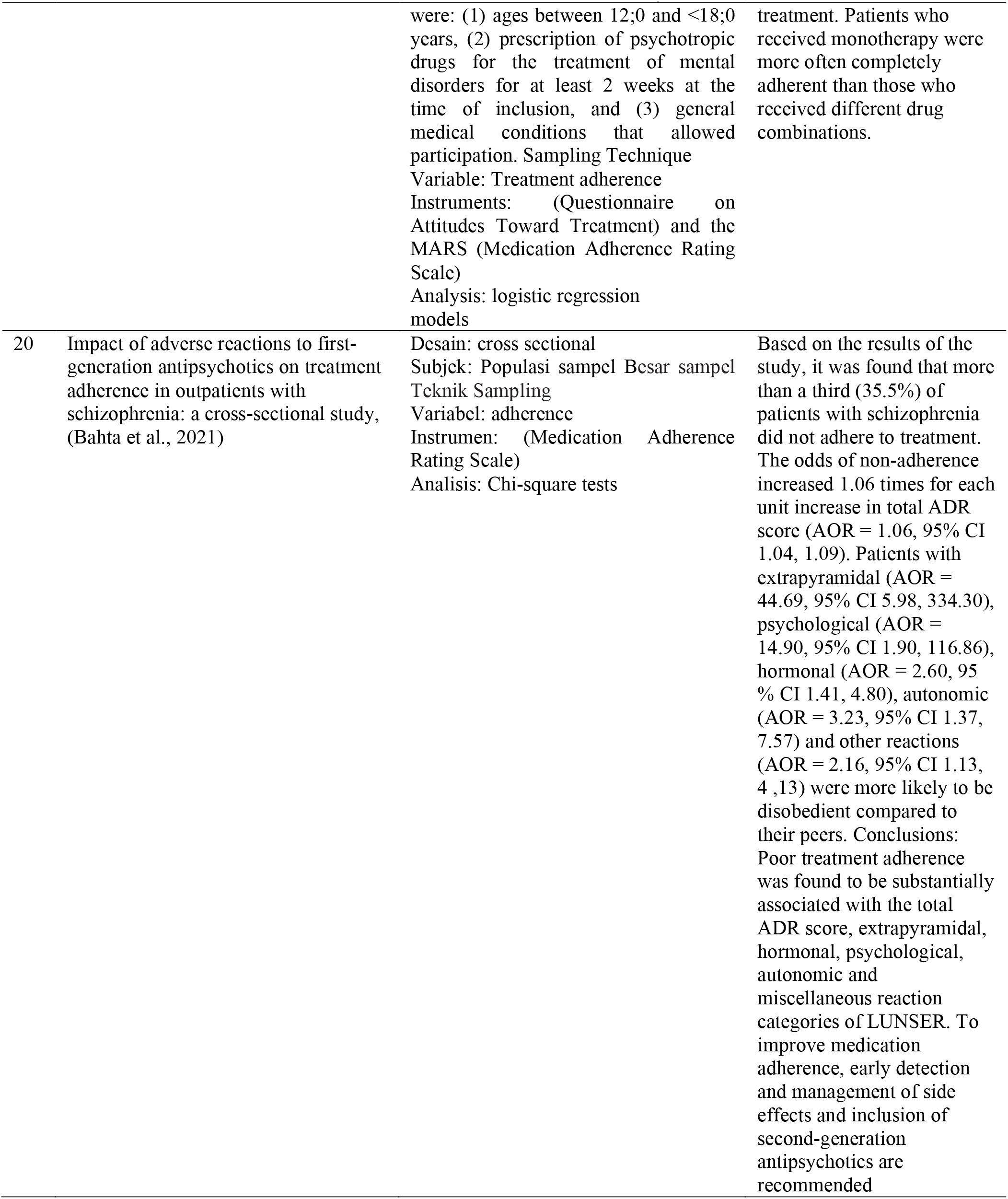
Summary of included studies

**Fig. 1.**
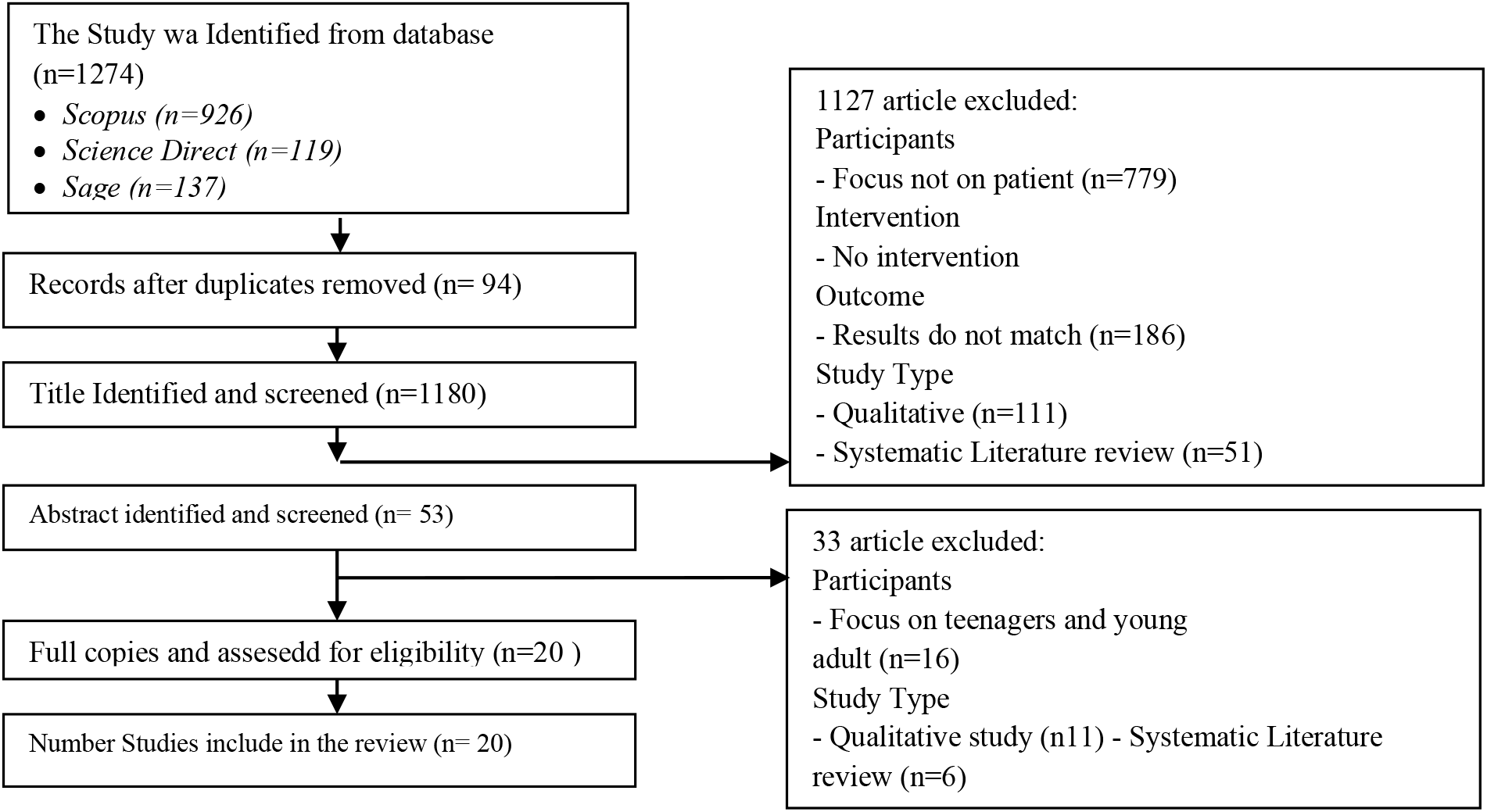
PRISMA diagram adapted from Moher et al. (2009).

## RESULT

Adherencee can be identified as the patient’s behavior towards the recommendations that have been given and has also been agreed between the patient and health workers, one of which is adherencee in taking medication. In this systematic review, treatment adherencee factors are classified into 2, namely:

1. Internal factors that affect medication adherencee in schizophrenic patients consist of (1) the patient’s own belief that medication is not needed for healing (2) relatively older age (3) self-efficacy (4) general health status (5) gender men (6) poorer treatment attitude (7) poor decision-making ability (8) poor insight (10) side effects
2. External factors External factors that affect medication adherencee consist of (1) lower socioeconomic status (2) living alone (3) length of illness (4) drug abuse (5) education level (6) severity of illness for example being in the acute phase (7) participating in mental health services (8) health facilities (9) marital status (10) receiving monotherapy
3. Psychosocial factors psychosocial factors that affect medication adherencee consist of (1) social support, (2) living with family

## DISCUSSION

This review aimed to analyze empirical findings that identified factors associated with medication adherence among adults with schizophrenia. The findings revealed two factors that are associated with medication adherence. Factors include Greater awareness of illness (insight), previous history of medication adherence, positive attitude toward medication, types of atypical antipsychotics, less severe psychotic symptoms, and social support, self-efficacy, general health status, gender men, lower socioeconomic status, living alone, length of illness, drug abuse, education level, severity of illness for example being in the acute phase, participating in mental health services, health facilities, marital status, receiving monotherapy.

Awareness of illness among schizophrenic patients is widely associated with two theories. Lack of awareness may be caused by psychological defense mechanisms as a form of refusal to face the perceived illness, or the presence of cognitive disorders that prevent them from understanding their illness better (Amador et al., 1991).

Previous history of medication adherence related to adherence can be explained through the Transtheoretical Model (Prochaska, DiClemente, & Norcross, 1992). This model suggests that each individual goes through stages of behavior change starting from pre-contemplation, contemplation, preparation, action and then the maintenance stage (Prochaska et al., 1992). Patients with adherence behavior are already in the maintenance phase which aims to prevent recurrence of the disease and enjoy the benefits of being obedient. Therefore, it is very likely that these patients will adhere to the next treatment period or the period of undergoing treatment.

Attitudes toward treatment stem from the perceived benefits of using drugs (Goodman, Knoll, Isakov, & Silver, 2005). In addition, negative attitudes are also associated with impaired working memory which may prevent patients from experiencing the real benefits of taking medication (Goodman et al., 2005). Therefore, those who have severe symptoms cannot make the right decision to take medication, which leads to non-adherence to taking medication. Social support consists of physiological and behavioral mechanisms, Through physiological mechanisms, social support activates the autonomic nervous system and the neuroendocrine system, which can help a person to reduce Through behavioral mechanisms, social support can influence health behavior. Therefore, social support must provide benefits for individuals such as positive health behaviors (adherence to taking medication) (Cohen, 1988).

Based on studies that have been conducted on patients with schizophrenia who are generally clinically stable, it was found that there is a correlation between insight in patients with medication adherence, the insight referred to in this case is clinical insight and insight into the knowledge possessed by the patient, (Lui et al., 2021). Knowledge can be a very effective motivator or reinforcer for someone, especially in this case, people with schizophrenia who do have a relatively longer time of onset of illness to learn to gradually change their behavior for the better, especially regarding adherencee to the treatment program that is followed. The attitude factor is also one of the patient’s internal factors that affect adherencee. Based on several research results, it was found that a poor or uncooperative attitude towards treatment is one of the factors causing non-adherencee to treatment undertaken by people with schizophrenia, (Sajatovic et al., 2021). Other influencing factors are age, fixed income, and disease severity and being in the acute phase of the disease has a significant effect on age medication adherencee. The elderly have the motivation to live healthy and always pay attention to their health, (Wang et al., 2020). The tendency to increase or increase in age is essential to improve medication adherencee, (Stockbridge et al., 2021); (Aylaz & Kılınç, 2017).

The side effects that arise are often the reason for non-adherencee to treatment undertaken by people with schizophrenia, extrapyramidal syndrome effects, hormonal effects, psychological effects on mood changes or other categories of reactions (Bahta et al., 2021). Side effects of treatment will also increase along with the multiple treatments that have been undertaken, patients who receive monotherapy or one type of drug are more often completely adhered to than those who receive different drug combinations, (Both et al., 2021). This is because the perceived side effects have an impact on the activities or daily life of people with schizophrenia, (Yu et al., 2021b). In some patients who receive different types of antipsychotics, they will get different responses or results so that this will also be one of the determinants of treatment adherencee, (Lee et al., 2019).

In a broader perspective, the attitude taken by the individual is the main and essential thing in determining the feeling or adherencee to the treatment being undertaken, (Lee et al., 2019). Individual attitudes in complying with all therapeutic regimens undertaken by various factors including their spiritual beliefs, based on the results of this study it is illustrated that there is a significant relationship between spiritual well-being and medication adherencee in individuals diagnosed with mental disorders, (Gültekin & Kavak Budak, 2021).

Family, social or external environment related to medication adherence in people with schizophrenia. In some studies it is described that patients with schizophrenia do not adhere to medication. length of hospitalization, health facilities, knowledge, medical social support, and motivation to use drugs, all of these factors are external factors outside the patient so that in principle there is a need to optimize this resource, (Hsieh et al., 2019).

### Implications of This Review

Findings from the results of this literature review can help design interventions based on: the factors that have been identified. Based on the information gathered, nurses can play a role in improving medication adherence among their schizophrenic patients in clinical practice. Awareness of the disease is closely related to medication adherence. Nurses can provide psychoeducation to clients and their families about their illness by increasing knowledge about illness and treatment (Sajatovic et al., 2021). Psychoeducation sessions should include elements such as symptom management. The presence of psychotic symptoms is also a factor associated with adherence shown in the literature. Because drug side effects are related to adherence and have been reviewed frequently, nurses should play an important role in monitoring drug use and reviewing drug side effects and discussing with patients strategies to improve them. side effects (Leijala et al., 2021).

## STUDY LIMITATION

There are several research limitations that need to be considered in this systematic review. First, there are thirteen articles (19.7%) which have almost the same research results but different research locations so that from the thirteen articles the identification results obtained are approximately the same. In addition, most of the studies (90.1%) were studies with a cross-sectional design so that it was not possible to determine a causal relationship. Apart from the limitations in this study, the articles discussed in the study were quite numerous (total articles = and varied so that the factors identified in this literature review can be considered as internal factors as well as external factors that affect medication adherencee.

## CONCLUSION

Overall, this review analyzed empirical findings that identified factors which are associated with medication adherence. These factors are important to design interventions to improve adherence. Literature gaps were identified, and suggestions for future interventions and research were also presented.

## Data Availability

All data produced in the present work are contained in the manuscript

